# Weather variables impact on COVID-19 incidence

**DOI:** 10.1101/2020.06.08.20125377

**Authors:** Javier G. Corripio, Lorna Raso

## Abstract

We test the hypothesis of COVID-19 contagion being influenced by meteorological parameters such as temperature or humidity. We analysed data at high spatial resolution (regions in Italy and counties in the USA) and found that while at low resolution this might seem the case, at higher resolution no correlation is found. Our results are consistent with a poor outdoors transmission of the disease. However, a possible indirect correlation between good weather and a decrease in disease spread may occur, as people spend longer time outdoors.

## Introduction

Weather impact on the transmission of SARS-CoV-2 virus causing the current COVID-19 pandemic is of interest to determine whether the infection might be seasonal. This could help contagion forecast models and be useful to mange health care resources. It is an area of active current research. Promising results were published at the initial stages of disease dispersion, showing graphically the correlation between the disease outbreak and narrow bands of air temperature or absolute humidity [1] as shown in Fig. 1. If the relationship between weather parameters and disease transmission could be established in a clear way, it would be a useful forecasting tool. Our aim is to evaluate if the relationship is maintained for a longer period of time, on a wider range of weather conditions and at a higher spatial resolution.

**Fig 1.**
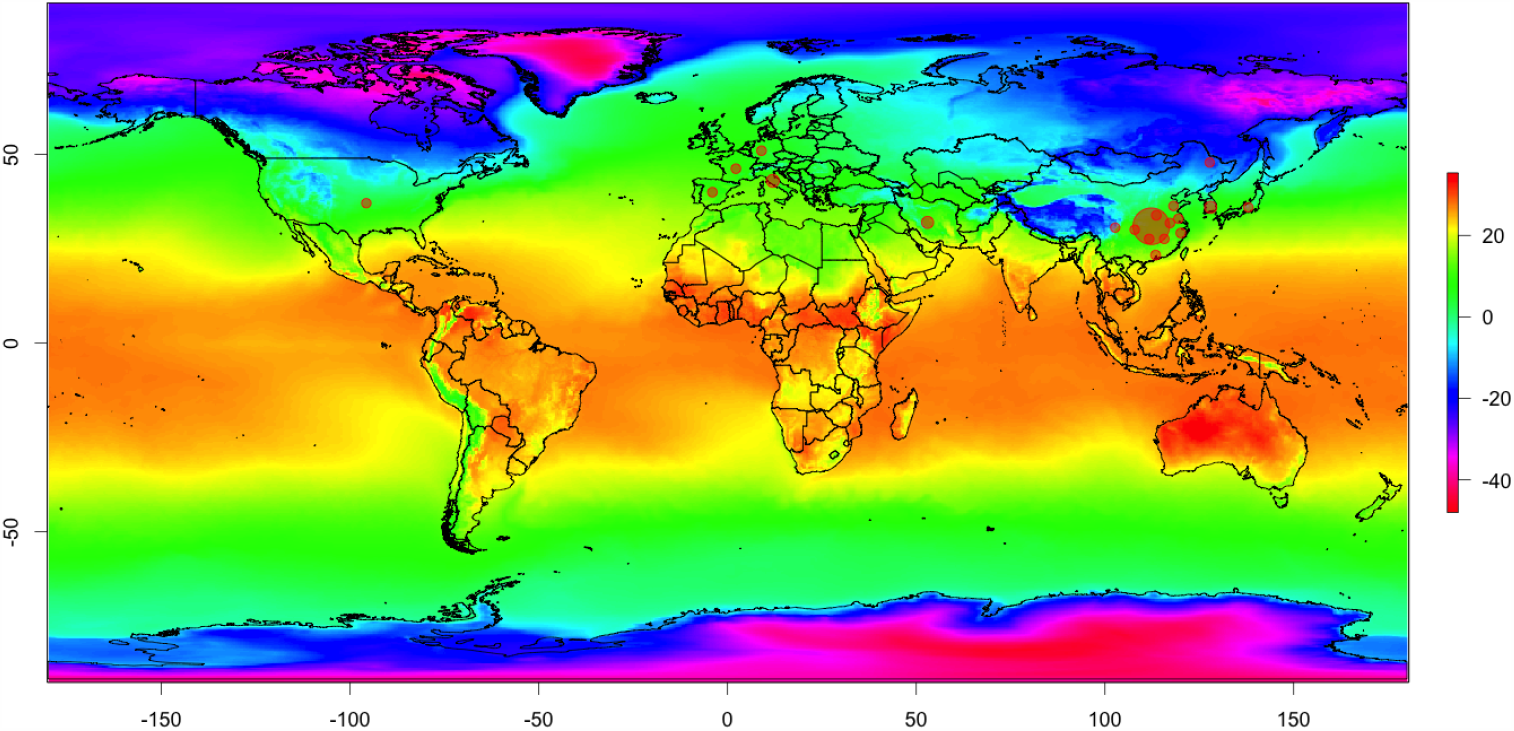
World temperature and cases. World average temperature [°C] during February and location of twenty spots with the higher number of COVID19 cases at the beginning of March, 2020. All spots are within a similar range of mean temperature except for Heilongjiang in northern China.

The relationship between meteorological parameters and disease spread has been investigated more amply in relation to the influenza outbreaks. In temperate regions influenza outbreaks show marked seasonality with an onset in autumn that continues through the winter. The cause of this seasonality is not fully understood. Travel [2], absolute humidity [3, 4] or school calendar [5] all appear to affect the spread of influenza, but the mechanisms behind these relationships are still unknown. Whereas other respiratory viruses seem to lack seasonality or peak in spring or summer, coronavirus outbreaks tend to begin in winter and end before the summer [6, 7].

Our intention is to find a simple meteorological model that can indicate whether meteorological parameters have any impact on transmission rates. We analyse the incidence data at high spatial resolution, for 21 regions in Italy and for 3142 counties in the USA, with weather data at comparable resolution interpolated to the center of every region or county.

## Materials and methods

### Meteorological data

Meteorological data were obtained from the NCEP GDAS/FNL 0.25 Degree Global Tropospheric Analyses and Forecast Grids [8] and the hourly ERA5 reanalysis from the European Centre for Medium Range Weather Forecasts ECMWF [9]. We compared both datasets to check if discrepancies are high. On average there is less than one degree centigrade difference for the daily average temperature and less than 1g/kg in specific humidity. As the ERA5 has higher temporal resolution, we used this data set for the statistical analysis.

Both ERA5 and GDAS provide the data in grib format (GRIdded Binary), that was processed using the Grid Analysis and Display System (GrADS) software [10]. Data were interpolated for every county and region coordinates from the beginning of January to the seventh of April, two weeks after lockdown was declared in many US states. This time frame was used so that both in Italy and the USA the window between the first cases and lockdown was analysed. In this way it is possible to compare spread before any lockdown effects. Italy went into lockdown on the ninth of March and USA between the 19th and the 24th of March. The resulting csv files with meteorological data are available with the supplementary material at https://meteoexploration.com/covid19/.

ERA5 does not provide specific or absolute humidity, but dew point temperature. From this variable and the local pressure we derived first water vapor pressure, following Bolton (Eq. 10) [11], and then specific and absolute humidity following Stull (Eq. 4.7 and 4.10) [12].

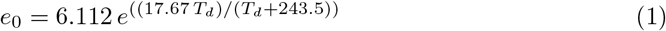

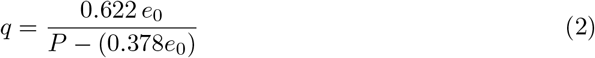

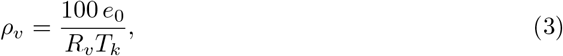

where *e*_0_ is actual water vapour pressure, *T*_*d*_ is dew point temperature in degrees centigrade, *q* is specific humidity in kg/kg, *P* is local atmospheric pressure in hPa, 0.622 = 18.016*/*28.966 is the ratio of molecular weights of water and dry air, *ρ*_*v*_ is absolute humidity, *R*_*v*_ is the gas constant for water vapor (461*JK*^−^1*kg*^−^1) and *T*_*k*_ is air temperature in Kelvin.

Different studies use either specific or absolute humidity, but the differences in the range of temperature and pressures found in this study are small. For the whole of Italy, for example, the mean difference is smaller than 1g of water vapour and values run almost parallel to each other.

Hourly data was aggregated as daily means and later aggregated as the mean over a period of eight days with a lag of two days with respect to incidence cases. The median incubation period of COVID-19 has been estimated to be 5.1 days, with 97.5% of those developing symptoms doing so within 11.5 days [13]. Thus, we tested a range of days and lags from seven to 14 and two to five respectively. We imposed a minimum of seven days for the accumulated incidence to avoid weekend errors in the data reporting. The final figures of eight and two gave the best result in the regression analysis.

### Epidemiological data

We retrieved global data on cases and mortality from the COVID-19 Data Repository by the Center for Systems Science and Engineering (CSSE) at Johns Hopkins University [14] and from the European Centre for Disease Prevention and Control [15]. Both datasets are consistent and updated daily. Italian data at regional level were downloaded from the github repository of the *Sito del Dipartimento della Protezione Civile* [16]. Data for the USA at county level were also obtained from the Johns Hopkins University repository. An additional attempt was made to use data from Spain, a country that has been heavily impacted by the pandemic. There are regional data for Spain, but this is too coarse a resolution, a single region such as Andalucía is larger than several European countries. Provincial data are very fragmented, without a single central repository. A crowdsourced effort to compile these data [17] has been made, but unfortunately the result has too many gaps.

### Geographical and other data

Global and European shapefiles are from Natural Earth [18]. Initially we used the taxpayer funded Eurostat GISCO: Geographical Information and maps - NUTS, but data usage is too restrictive while Natural Earth is completely open. Shapefiles for the USA were downloaded from the TIGER/Line Shapefiles from the United States Census Bureau [19]. Data for the population of Italian regions were obtained from the Italian *Istat - Istituto nazionale di statistica* [20]. For US county population, data were obtained from the United States Census Bureau [21]. Additional, non conclusive tests were done with airport transportation data, that were obtained from the EU Open Data Portal [22] and the Federal Aviation Administration [23].

### Statistical analysis

A first step is using descriptive statistics and spatial visualisation that can help identifying any possible correlation between weather and disease transmission. A second step is designed following Barreca and Shimshack [24]. They found that half of the average seasonal differences in US influenza mortality can be explained by seasonal differences in humidity alone, and there seems to be an important threshold at 6g/kg. Because mortality is dependant not only on contagion, but also on the health system capacity and resources, we used the number of cases accumulated over a period of time. Following Barreca and Shimshack we asume that incidence can be simulated with a simple statistical model as:

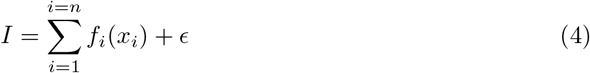

where *f*_*i*_(*x*_*i*_) is a function of variable *x*_*i*_ and *E* a general error term.

This model was implemented in the R statistical package using generalised linear models (glm) [25] and generalised additive models (gam) from the *mgcv* library [26]. We are interested in response to meteorological variables, but we also tested population density and passenger emplainment at nearest airport with very poor results.

## Results and discussion

The visualisation of Italian data in Figures 2 to 4 seems to confirm the hypothesis that transmission is hindered by specific humidity above 6g/kg and mean air temperature above 11°C [1]. Fig. 2 shows the evolution of accumulated incidence during eight days in Italy from the beginning of February to the 23rd of March, two weeks after general lockdown. Every colour represents one region and the points increase in size with the date, the name of the region is plotted at the last date. It appears that regions with specific humidity higher than 6 g/kg of water vapour at the beginning of the period did not develop many cases. Similarly in Fig. 3, a threshold slightly over 11°C seems to hinder the development of cases. Fig. 4 shows the mean ultraviolet radiation, and it appears that regions with values higher than 18 Wm^−2^ at the beginning of the episode remain at very low incidence. Valle d’Aosta seems to be an exception, but the increase in UV radiation is towards the end, while at the beginning of the period values were below the apparent threshold. This is a small region at high altitude in the Alps, bordering with Switzerland, where clear days in March brought about a high dose of UV radiation.

**Fig 2.**
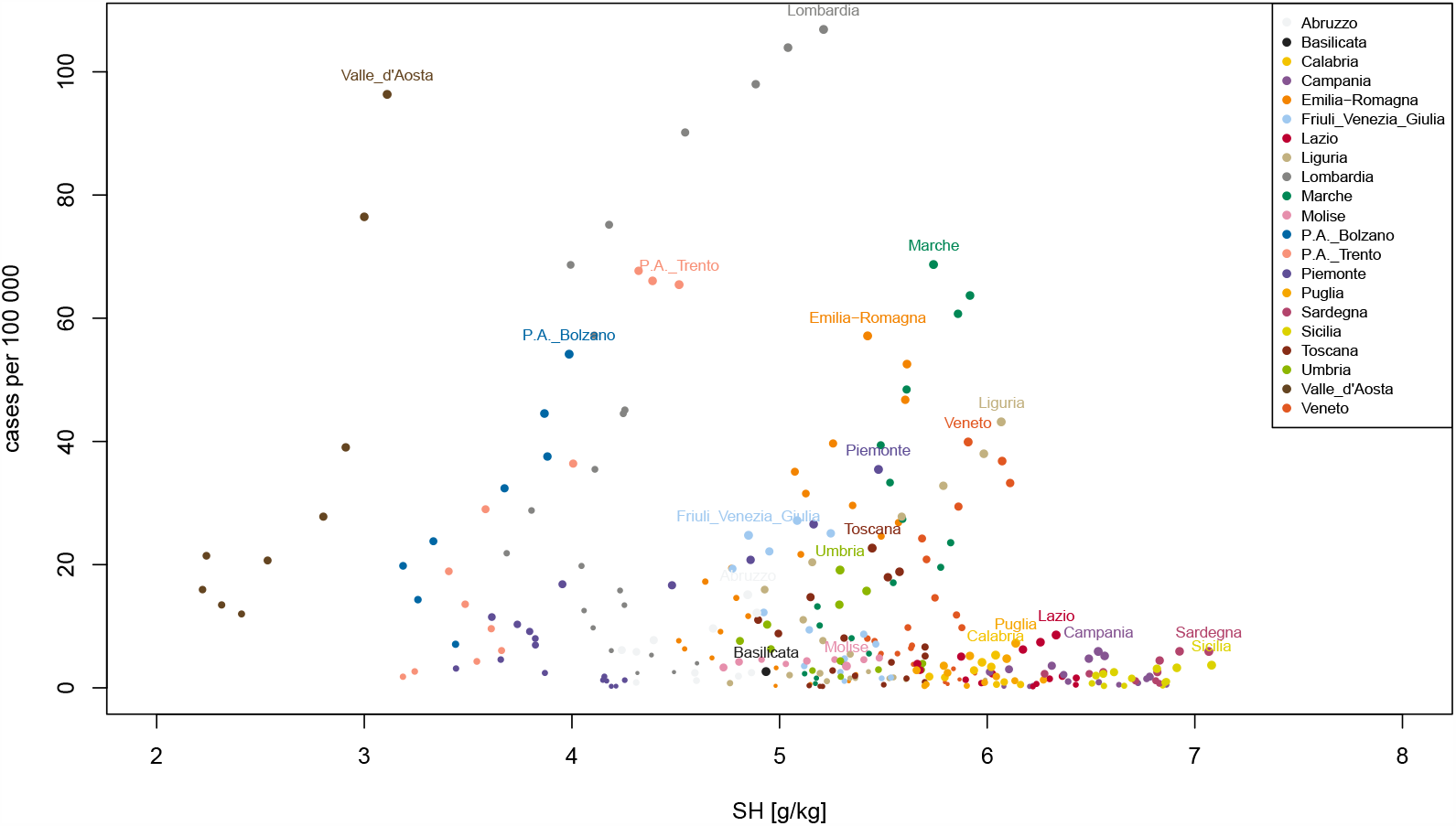
Evolution of cases in Italy and specific humidity. Cases per region in Italy against specific humidity. Every dot colour corresponds to an Italian region, size of dots increase with time from the 11th of February to the 23rd of March, two weeks after Italy’s lockdown.

**Fig 3.**
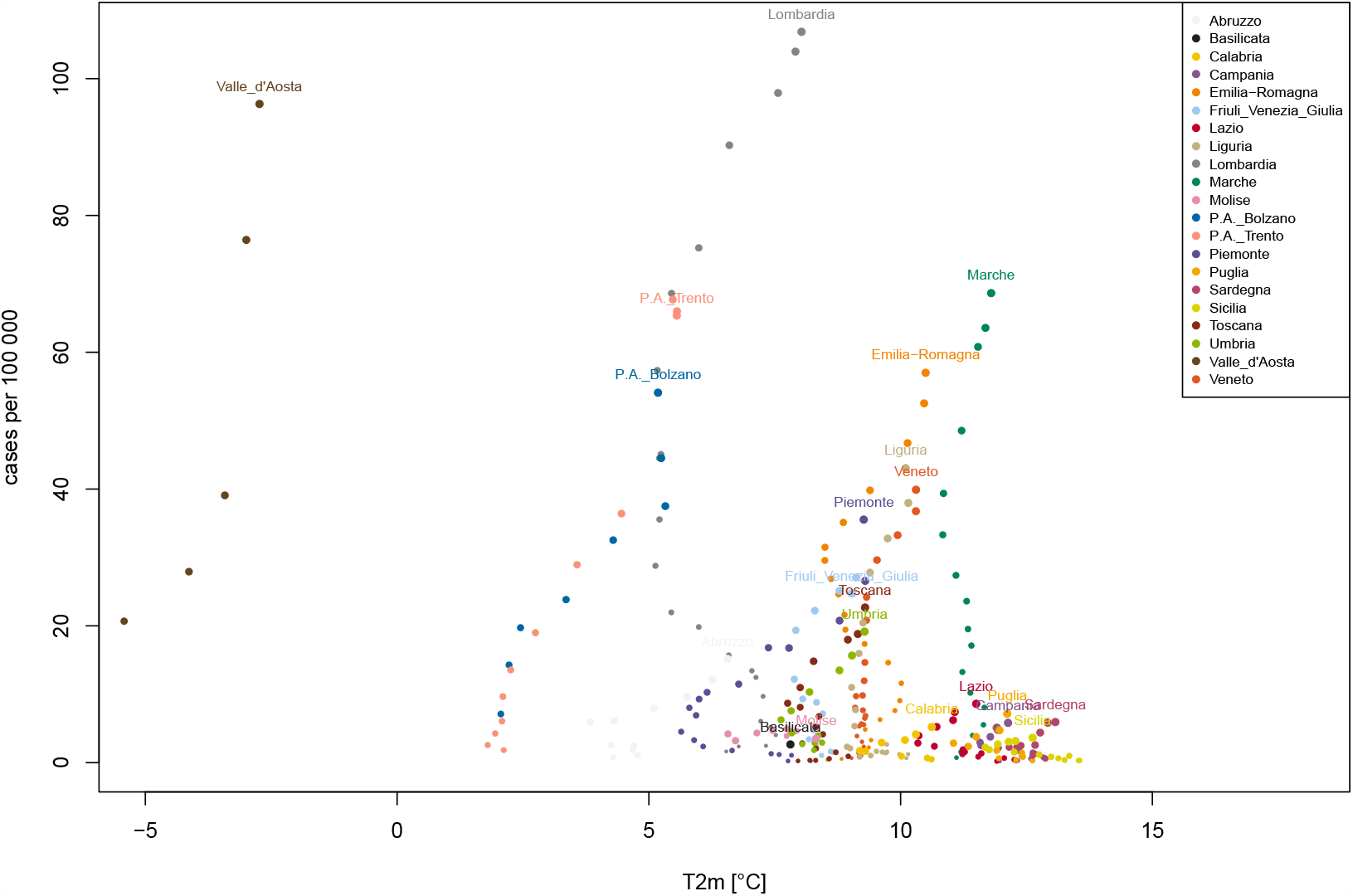
Evolution of cases in Italy and temperature. Cases per region in Italy against air temperature. Every dot colour corresponds to an Italian region, size of dots increase with time from the 11th of February to the 23rd of March, two weeks after Italy’s lockdown.

**Fig 4.**
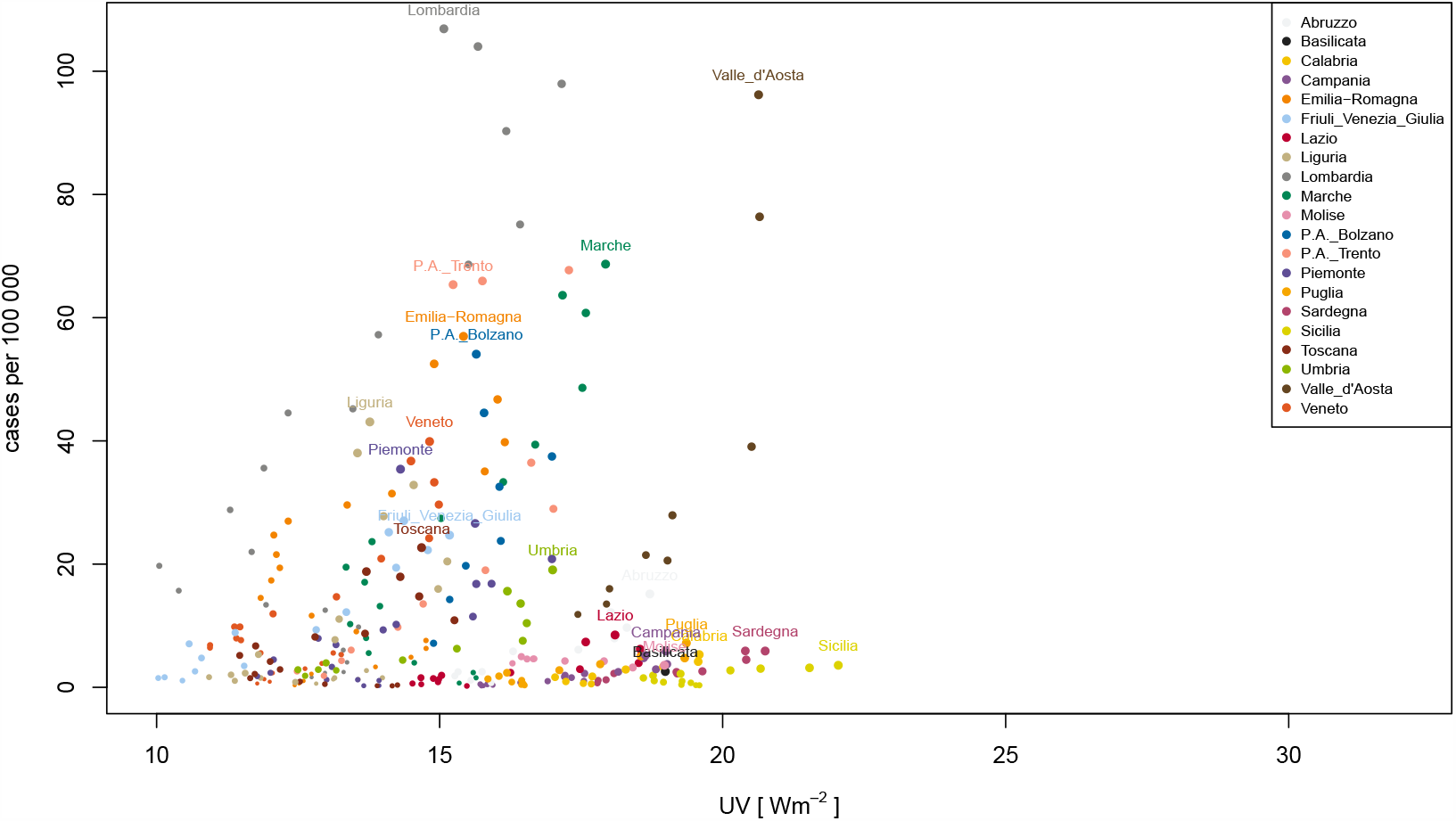
Evolution of cases in Italy and UV radiation. Cases per region in Italy against ultraviolet radiation. Every dot colour corresponds to an Italian region, size of dots increase with time from the 11th of February to the 23rd of March, two weeks after Italy’s lockdown.

Additional tests were done for precipitation, but as Fig. 5 shows there is no evident relationship, thus we discarded this variable for further analysis. A statistical model using generalised linear models with Poisson error structure indicate that all three parameters (specific humidity, temperature and ultraviolet radiation) are significant, but the results have a very low value of adjusted R-squared, as low as 0.14. A model with generalised additive models seems to explain 32% of the deviance, with a poor R-squared value of 0.18. We used count data, incidence accumulated over eight days for the models but the figures show incidence per 100 000 inhabitants to help the visualisation.

**Fig 5.**
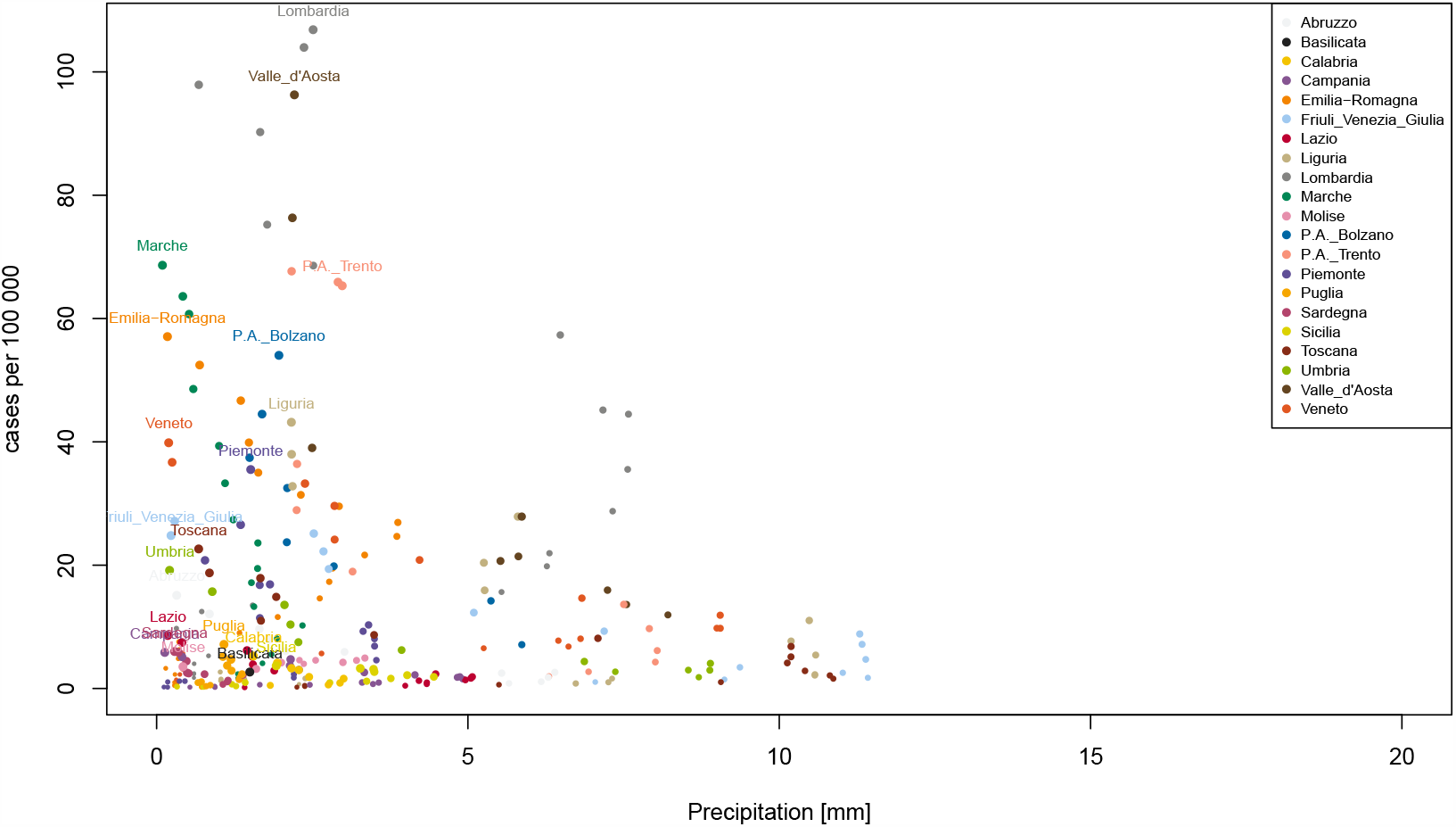
Evolution of cases in Italy and precipitation. Cases per region in Italy against precipitation. Every dot colour corresponds to an Italian region, size of dots increase with time from the 11th of February to the 23rd of March, two weeks after Italy’s lockdown.

The threshold hypothesis, which could be useful in forecasting future seasonal incidence, is refuted by the high resolution data from the USA. As can be noted from Figures 6 to 8, there is no evidence of thresholds whatsoever in these data. Fig. 6 shows incidence per capita with respect to Specific Humidity. There are too many counties to use distinctive colours, so these are depicted as a range from grey to blue according to the initial values. Although there seems to be some clustering, especially at high values, there is a broad dispersion at lower values for most of the points. The range of the dispersion is limited by the maximum mean temperature at every county at this period of the year, as maximum possible specific humidity increases exponentially with temperature. Blaine, New York, Rockland and Westchester counties are outside the chart. The statistical modelling with glm results in a very low R squared value of 0.02 and only temperature seems to be significant, but this is probably a spurious correlation, as the number of cases increase from winter towards the spring, which correspond to a seasonal increase in temperature.

**Fig 6.**
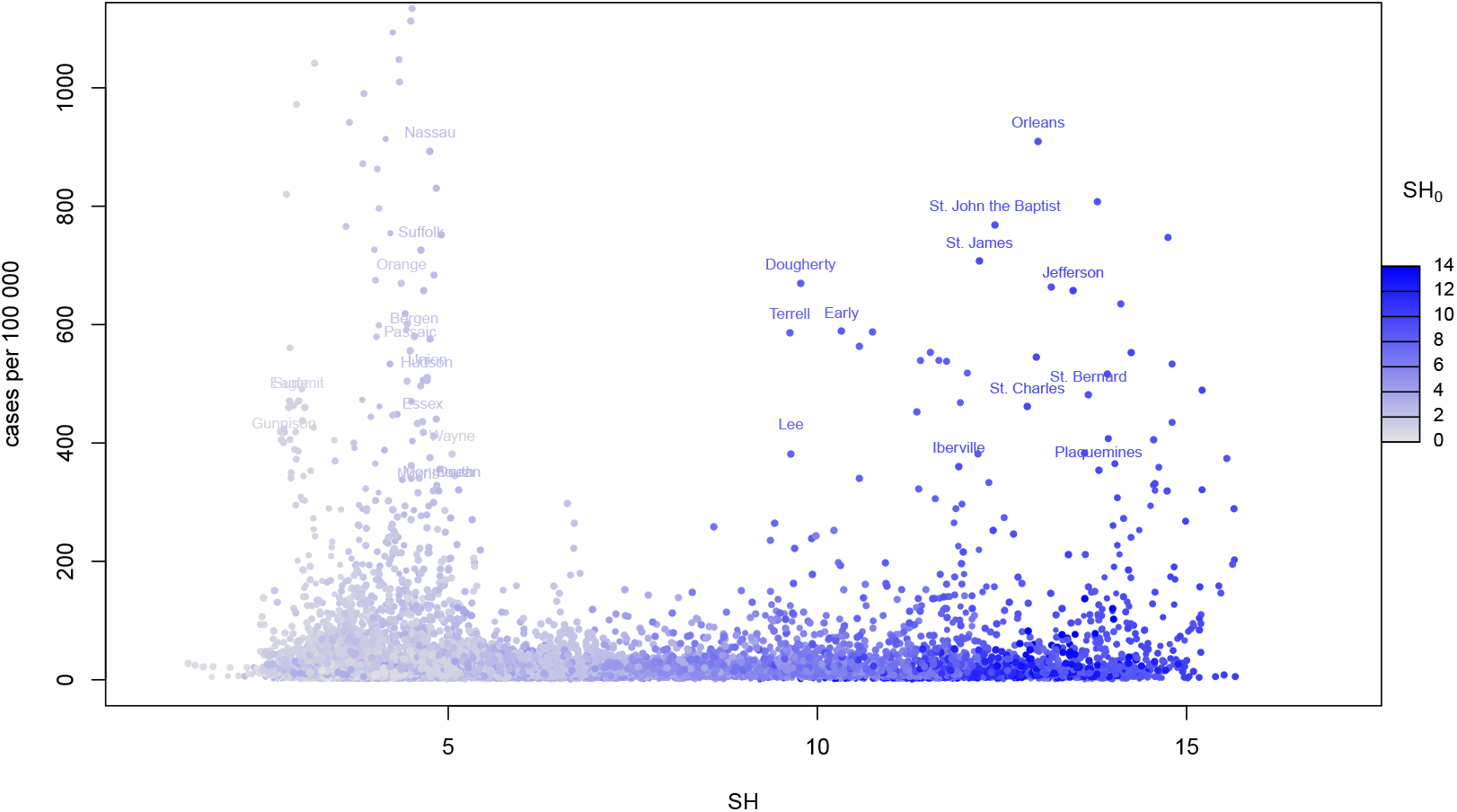
Evolution of cases in the USA and specific humidity. Cases per county in the USA against specific humidity. Dot colours are graded from grey to blue according to initial values of specific humidity (low to high), size of dots increase with time from the 20th of February to the seventh of April, between 10 days to two weeks after lockdown depending on State.

**Fig 7.**
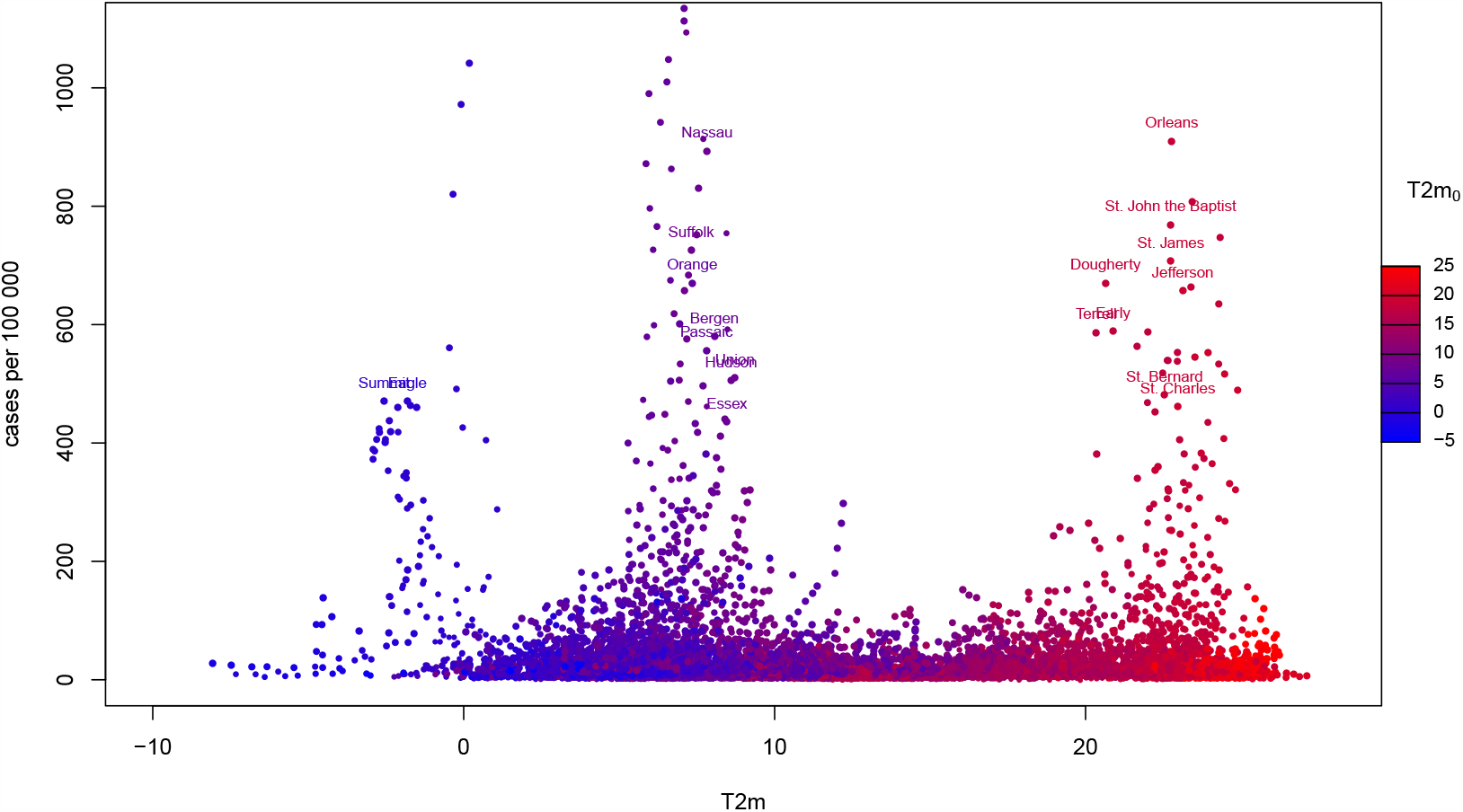
Evolution of cases in the USA and temperature. Cases per county in the USA against air temperature. Dot colours are graded from blue to red according to initial values of air temperature (low to high), size of dots increase with time from the 20th of February to the seventh of April, between 10 days to two weeks after lockdown depending on State.

**Fig 8.**
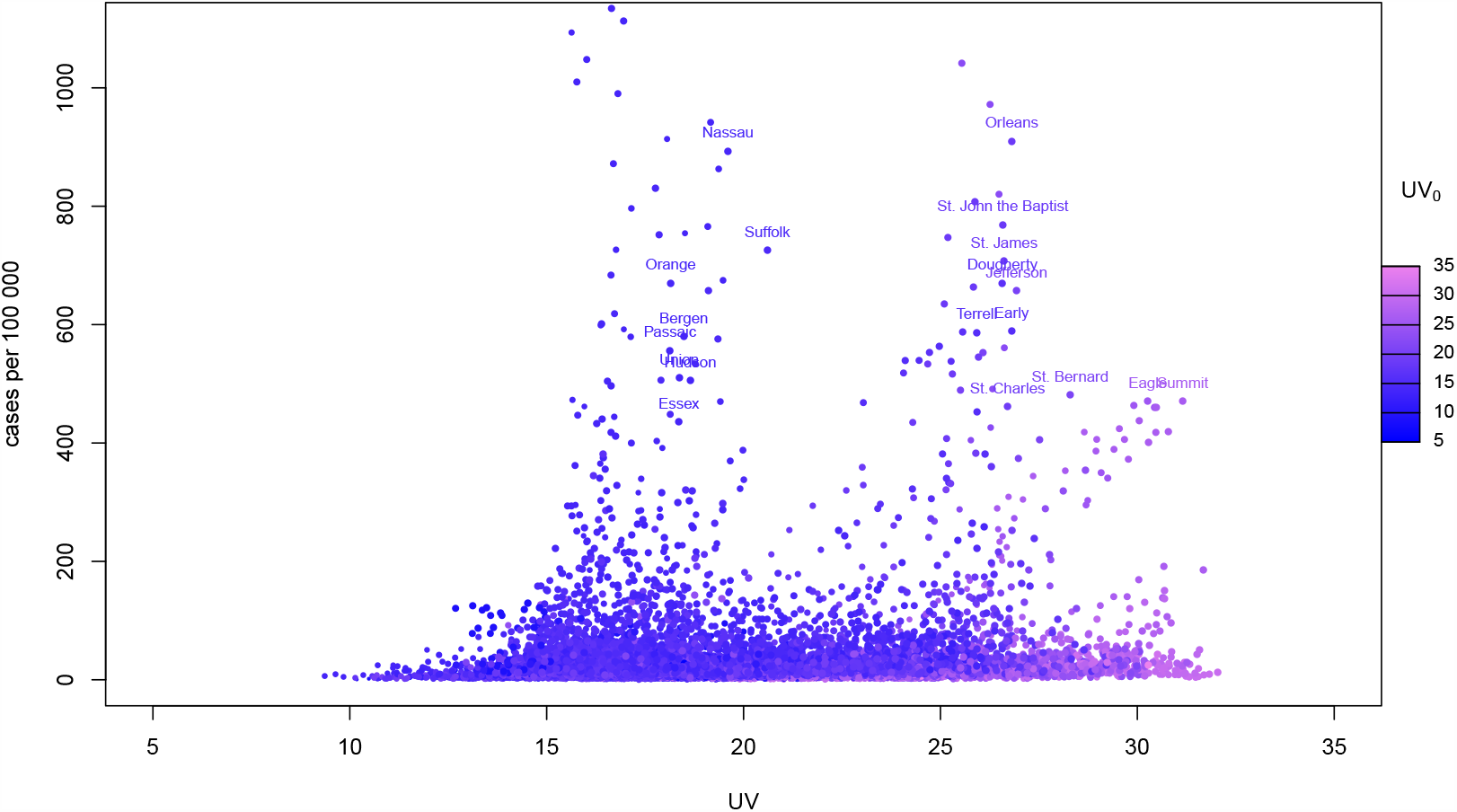
Evolution of cases in the USA and UV. Cases per county in the USA against ultraviolet radiation. Dot colours are graded from blue to violet according to initial values of ultraviolet radiation (low to high), size of dots increase with time from the 20th of February to the seventh of April, between 10 days to two weeks after lockdown depending on State.

Our results suggest that the spread of SARS-CoV-2 is not influenced by outdoors specific humidity, temperature or UV radiation. If we consider some of the literature on influenza spread, several experiments show that in closed spaces increased humidity reduces the influenza virus droplet transmissions [27–29]. The mechanics behind this effect has been explored by Yang and Marr [30]. Kormuth et al. [31] found this is probably true only for droplet transmission as they confirmed aerosol transmission to be the same even at high relative humidity. However, there does not seem to be much literature on contagion rates in the open air.

Jaakkola et al. [32] found that changes in temperature and absolute humidity were more likely to cause infection in Finnish military conscripts. They suggest this may be due to cold stress and a change in the susceptibility of the air passage to viral infections. This is consistent with Davey and Ried [33] who also found an increase in influenza cases with a decrease in temperature in the UK, a finding supported by Davies et al. [34] who correlated increased pulmonary disease mortality rates with unusually cold and dry weather spells. Willem et al. [35] find that part of this weather effect may be behavioural. As with warmer temperatures people spend more time outside, therefore decreasing the contact with people in closed spaces. This may go some way to explaining the differences between USA and Italy. In Italy much of the spread was late winter whereas in the USA it was already the beginning of Spring, when days are getting longer and people spend more time outside. It is also a plausible explanation for Italy, where most of the cases were concentrated in the Po Valley. The Po is often cold and cloudy from autumn to spring, a weather less inviting to outdoor life than in the southern regions, where clear skies and moderate cold are common even in winter. A poor outdoors contagion is also evidenced by the high rate of contagion in some ski resorts of the Alps, such As Valle d’Aosta in Italy or Ischgl and Solden in Tirol, where most of the cases can be traced back to just a few aprés ski centres [36].

## Conclusion

Although initial data at coarse resolution seems to suggest a narrow range of meteorological conditions for the spread of COVID-19, a closer look at higher resolution does not support this hypothesis. Higher resolution data reveal no correlation between meteorological parameters and COVID-19 incidence. This lack of correlation may be indicative of a poor outdoor transmission. It could be that lower outdoor transmissions have an indirect relationship with meteorological parameters. The good weather and longer days, will affect behaviour as people tend to spend more time outdoors, making the spread of the disease less likely. It may also explain the differences in contagion rates between the north and the south of Italy, due mostly to behavioural differences, and it is supported by the high relative importance of a few “hot spots” in the spreading of the disease, in agreement with Endo’s work [37].

## Supporting information

### S1 Data availability

Meteorological data interpolated to the coordinates of every region in Italy and every county in the USA, figures at higher resolution and additional figures are available at https://meteoexploration.com/covid19/.

## Data Availability

All data used in this study are available, a link is provided for downloading them

https://meteoexploration.com/covid19/

## Acknowledgments

We are grateful to the different Agencies, Research Centres and Universities that have made data readily available. meteoexploration.com provided computer resources and staff time. We did not receive any funding for this work.

